# Using a hybrid of artificial intelligence and template-based method in automatic item generation to create multiple-choice questions in medical education: Hybrid AIG

**DOI:** 10.1101/2024.07.15.24310424

**Authors:** Yavuz Selim Kıyak, Andrzej A. Kononowicz

## Abstract

**Objectives:** Template-based automatic item generation (AIG) is more efficient than traditional item writing but it still heavily relies on expert effort in model development. While non-template-based AIG, leveraging artificial intelligence (AI), offers efficiency, it faces accuracy challenges. We aimed to integrate these approaches for leading to a significant rise in efficiency for AIG without sacrificing accuracy.

**Material and Methods:** We proposed the Hybrid AIG method that utilizes AI to generate item models (templates) and cognitive models to combine the advantages of the two AIG approaches. The Hybrid AIG consists of seven steps. The first five steps are carried out by an expert in a customized AI environment. Following a final expert review (Step 6), the content in the template can be used for item generation through a traditional (non-AI) software (Step 7). We used two multiple-choice questions for demonstration.

**Results:** We demonstrated that AI is capable of generating item models and cognitive models for AIG under the guidance of a human expert. Leveraging AI in template development has substantially reduced the time investment from five hours to less than 10 minutes, and made it significantly less challenging.

**Conclusions:** The Hybrid AIG method transcends the traditional template-based approach by marrying the “art” that comes from AI as a “black box” with the “science” of algorithmic generation under the oversight of expert as a “marriage registrar”. It does not only capitalize on the strengths of both approaches but also mitigates their weaknesses, offering a human-AI collaboration to increase efficiency in medical education.

## Introduction

Multiple-choice items are integral to written assessment in medical education, serving as a useful tool for assessing a wide range of knowledge and skills. Their common use spans from evaluating factual knowledge to clinical reasoning and decision-making in various domains.^1^ This assessment format supports high-volume testing with the added advantage of automated scoring to enhance the efficiency of examinations in medical education.

Traditional way of writing multiple-choice items, characterized by manual development processes, presents significant challenges in scalability. This challenge stems from the intensive time and effort required to create and review each question. This laborious process, which demands expertise and resources, faces a bottleneck in scaling up to meet the escalating demand for a vast quantity of quality items. The challenge is particularly pronounced in medical education, where only a progress test administration in a year requires having 2400 multiple-choice items,^2^ showing the inefficiency of traditional methods in satisfying the needs of question banks in medical schools.

Template-based automatic item generation (AIG) is a scalable method used in educational assessment that leverages predefined templates to systematically produce multiple-choice items with the help of software.^3^ It has been implemented in six languages including Turkish.^3–5^ It consists of three sequential stages; development of a cognitive model, development of an item model (template), and using software for rapid generation of hundreds of items.^6^ Item models developed based on cognitive models are structured by subject matter experts to include variables and rules for item generation, allowing for a more efficient creation of consistent questions. This approach enhances the scalability of item development/review compared to traditional item writing,^3^ addressing the growing demand for high-quality assessment materials across various educational domains. Template-based AIG, while generating high-quality multiple-choice items efficiently,^7–11^ still heavily relies on expert effort in development of cognitive models and item models. Although it depends on the content area and the expert’s skills, development of one cognitive model takes three hours, and one item model takes two hours for a subject matter expert who is also experienced in template-based AIG.^6^ Moreover, this development process necessitates high levels of extraneous cognitive load due to high element interactivity^12^ that requires experts to deal with several components simultaneously. Therefore, “creating cognitive models for AIG is challenging”^3(p30)^ and it “requires a lot of practice” for experts^3(p30)^.

On the other hand, non-template-based AIG, which “can be guided by the syntactic, semantic, or sequential structure of a text”,^3(p57)^ is an approach that leverages natural language processing to generate assessment items without relying on predefined templates. Unlike the template-based method, this method utilizes the ability of artificial intelligence (AI) to produce content dynamically, for example, using ChatGPT, which is an AI-based chatbot developed by OpenAI, for creating items based on specific topics or learning outcomes provided by users.^13–17^ This approach allows for the generation of diverse and complex questions in seconds, offering flexibility and efficiency in item development. However, this AI-driven approach struggles with issues of inaccuracy and inconsistency,^15^ especially when good prompting strategies^18^ are not employed^19^. In AI-driven item generation, such as with ChatGPT, these issues often emerge due to the model’s reliance on its training data, which may not always align perfectly with the specific objectives intended by educators. For example, an AI might generate content that includes incorrect information, such as asserting that “the human heart only has two chambers”,^20^ or misinterpret the complexity level required for a medical education context. Furthermore, the “black box”^21^ nature of these AI models complicates diagnosing and correcting these errors within the AI mechanism, as it is challenging to trace back how the AI arrived at a particular output. Therefore, the process still requires subject matter experts to review and revise each generated question.^14,15,17,18,22^ Although it is more efficient than traditional item writing, necessity for reviewing each question is still inefficient.

As outlined above, recent advancements in AIG have offered efficiency, yet each method— template-based and non-template-based—brings its own set of limitations. The gap, therefore, lies in the need for a method that merges the structured efficiency of template-based AIG with the content generation capabilities of AI-driven, non-template-based, approaches. This convergence could potentially address the pressing need for tools augmenting capabilities of medical educators in test development. We are aware of the interdependence of social (human) and technical elements within an organization,^23^ advocating for the design of systems that concurrently optimize both human and technological components to achieve effective outcomes. In AIG, this can be interpreted as the need to harmonize the collaboration of a subject matter expert and AI tools working together on developing item and cognitive models.

In this paper, we propose a hybrid AIG method that utilizes AI to generate an item model (template) and a cognitive model for applying the item template in a template-based item generation process. This capitalizes on the strengths of both approaches but also mitigates their respective weaknesses, offering a novel human-AI collaboration to increase AIG efficiency in medical education.

## Methods and Results

This is a proof-of-concept study. As no human has participated in this study, there is no requirement for obtaining an ethical approval. We will present methods and results together in the same way that Gierl et al.^6^ described the template-based AIG.

The aim of the proposed Hybrid AIG method is to enhance the generation of multiple-choice questions by combining expert effort with AI. To optimize the collaboration between AI and experts in AIG system, we designed a new method where AI shoulders the significant cognitive load involved in template development, therefore reducing the cognitive burden on experts.

The Hybrid AIG consists of seven steps. The last two steps are carried out outside the AI environment. The AI environment necessitates a specialized AI trained for generating item models and cognitive models. We used GPT Builder, which is a platform tailored to customize ChatGPT according to the user’s needs,^24^ to train our Custom GPT, titled *Item Model Maker for AIG*, for this purpose. It is accessible from this link: https://chat.openai.com/g/g-ISoiQOLyv-item-model-maker-for-aig

### Step 1: Providing a parent item

The origin point of the item model and the cognitive model development in the Hybrid AIG is a parent item, as it is in the item model development in the template-based method. It functions as a prototype for generating new questions that outlines the structure.

In the first step of the Hybrid AIG, the expert provides a parent item to AI. Although it is not a requirement, it would be better if the parent item has been chosen from well-performed items in an exam. This could mitigate the possible problems that can stem from the lack of quality in the parent item. If an expert does not have a parent item, they can use AI tools, such as Case-Based MCQ Generator, to generate an MCQ as a parent item.^16^

The purpose of generating MCQs in medical education is for a human expert to assess the students’ skills and knowledge and to evaluate whether they have grasped the nuances of the information presented, aiming for outcomes that are consistent with expert reasoning. The parent item includes the necessary components and embedded possible cognitive models required for this purpose, as it is approved and provided by the expert. It serves as the well-established basis and acts as a seed for generating a new template. In the next steps, AI will primarily handle the cognitive work in utilizing one of the possible underlying cognitive models in the parent item to generate a template. The expert’s role will be to monitor and ensure the AI maintains a consistent cognitive model in its output.

In order to demonstrate, we used two items, one from Gierl et al.’s work,^3^ and one from Turkish National Medical Specialty Exam, TUS (2021/1, clinical question number: 58). The reason behind choosing Gierl et al.’s item is that it allows readers to compare it with the existing item model developed by a subject matter expert, and the reason behind choosing a TUS item is that Gierl et al.’s item model probably has already been read by ChatGPT, so we also aimed to focus on an item that was not modelled before. We presented the parent items provided to AI below.

Gierl et al.’s Item:

“A 22-year-old female sees her doctor and reports that she’s been experiencing a mild cough and slight body aches that have developed over a few days.

Upon examination, she presents with an oral temperature of 37°C. What is the most likely diagnosis?

A. Hay fever
B. Ear infection
C. Common cold
D. Acute sinusitis
E. Seasonal influenza”

TUS Item:

“A fifteen-year-old girl, who became ill during her physical education class after lunch, is brought to the infirmary complaining of coughing, shortness of breath, dizziness, facial swelling and redness, as well as itching in her hands and feet. During the physical examination, the patient appears anxious, with a blood pressure reading of 80/50 mmHg, periorbital edema, and wheezing upon auscultation. Which of the following is the most likely diagnosis for this patient?

A. Exercise-induced asthma
B. Cholinergic urticaria
C. Pulmonary embolism
D. Anaphylaxis
E. Vasovagal syncope”

### Step 2: Identification of elements for manipulation

In this step, expert identifies words or phrases in the parent item for variable manipulation.

In the identification process, the expert can, preferably, consider the importance of the content in the case that is more influential in deciding what the correct answer is. Table 1 shows AI’s response and what the expert chose as the elements for manipulation.

**Table 1.** Identification of elements for manipulation.

| Who speaks | Gierl et al.'s Item | TUS Item |
| --- | --- | --- |
| AI | Thank you for sharing the parent item. Could you please identify words or phrases in the question that you would like to manipulate for variable creation? These can include demographic details, symptoms, examination findings, or any other elements that could vary across different versions of this question. | Based on the parent item you've provided, let's proceed to the next step. Could you please identify words or phrases in the scenario and question that you would like to have as variable elements for manipulation? These could be specific symptoms, the patient's demographics, the setting, or any other details you think should be variable. |
| Expert | Element1: mild cough<br>Element2: slight body aches<br>Element3: over a few days<br>Element4: 37°C | Element1: during her physical education class after lunch<br>Element2: coughing, shortness of breath, dizziness, facial swelling and redness<br>Element3: a blood pressure reading of 80/50 mmHg, periorbital edema, and wheezing upon auscultation |

### Step 3: Replacement of elements and choosing options

In this step, AI replaces the words/phrases received from the expert with the elements in brackets, then the expert reviews the output, and prompts AI to make necessary changes, if any. Subsequently, the expert chooses two or more options from the options in parent item. The AI will use them to define the possible values of elements. Table 2 presents the replaced version and the expert’s choice among the options.

**Table 2.** Replacement of elements and choosing options.

| Who speaks | Gierl et al.'s Item | TUS Item |
| --- | --- | --- |
| AI | Based on the elements you've identified, here's how the parent item could be modified with these variables in brackets:<br>"A 22-year-old female sees her doctor and reports that she's been experiencing a [Element1] and [Element2] that have developed [Element3]. Upon examination, she presents with an oral temperature of [Element4]. What is the most likely diagnosis?<br>A. Hay fever<br>B. Ear infection<br>C. Common cold<br>D. Acute sinusitis<br>E. Seasonal influenza"<br>Could you please choose two or more options from the parent item that the variables will be focused | Great, I'll incorporate these elements as variables into the parent item. Here's how it will look:<br>"A fifteen-year-old girl, who became ill [Element1], is brought to the infirmary complaining of [Element2], as well as itching in her hands and feet. During the physical examination, the patient appears anxious, with [Element3]. Which of the following is the most likely diagnosis for this patient?<br>A. Exercise-induced asthma<br>B. Cholinergic urticaria<br>C. Pulmonary embolism<br>D. Anaphylaxis<br>E. Vasovagal syncope"<br>Now, could you please choose two or more options from the answer choices that you'd like the variables to significantly impact?<br>This will help in designing the variables for |
|  | on? This will help in offering variables for each element that significantly impact the correct answer according to medical sources and guidelines. | each element to ensure they align closely with the medical scenarios corresponding to the chosen options. |
| Expert | Common cold<br>Seasonal influenza | B. Cholinergic urticaria<br>D. Anaphylaxis |

### Step 4: Offering values in elements

In this step, AI offers possible values for each element that can impact the correct answer Then, the expert reviews the output and makes necessary changes by prompting, if any. Table 3 shows the response of AI in order for offering values. It is important to note that “values” were presented as “variables” by AI in the example because it did not work well when we call them “values” (e.g. it was prone to offer numbers rather than words). It may not be a problem in newer versions.

**Table 3.**
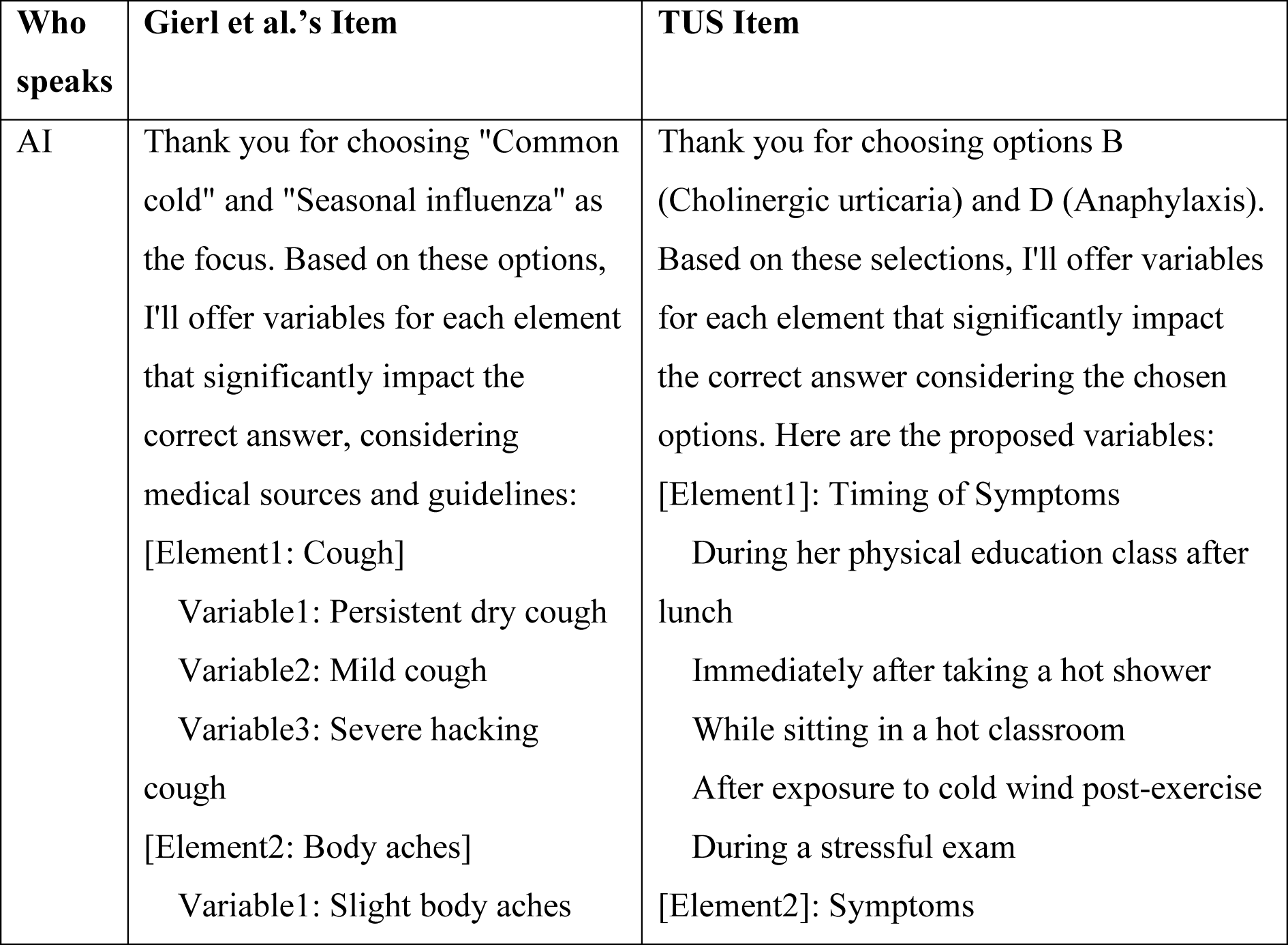
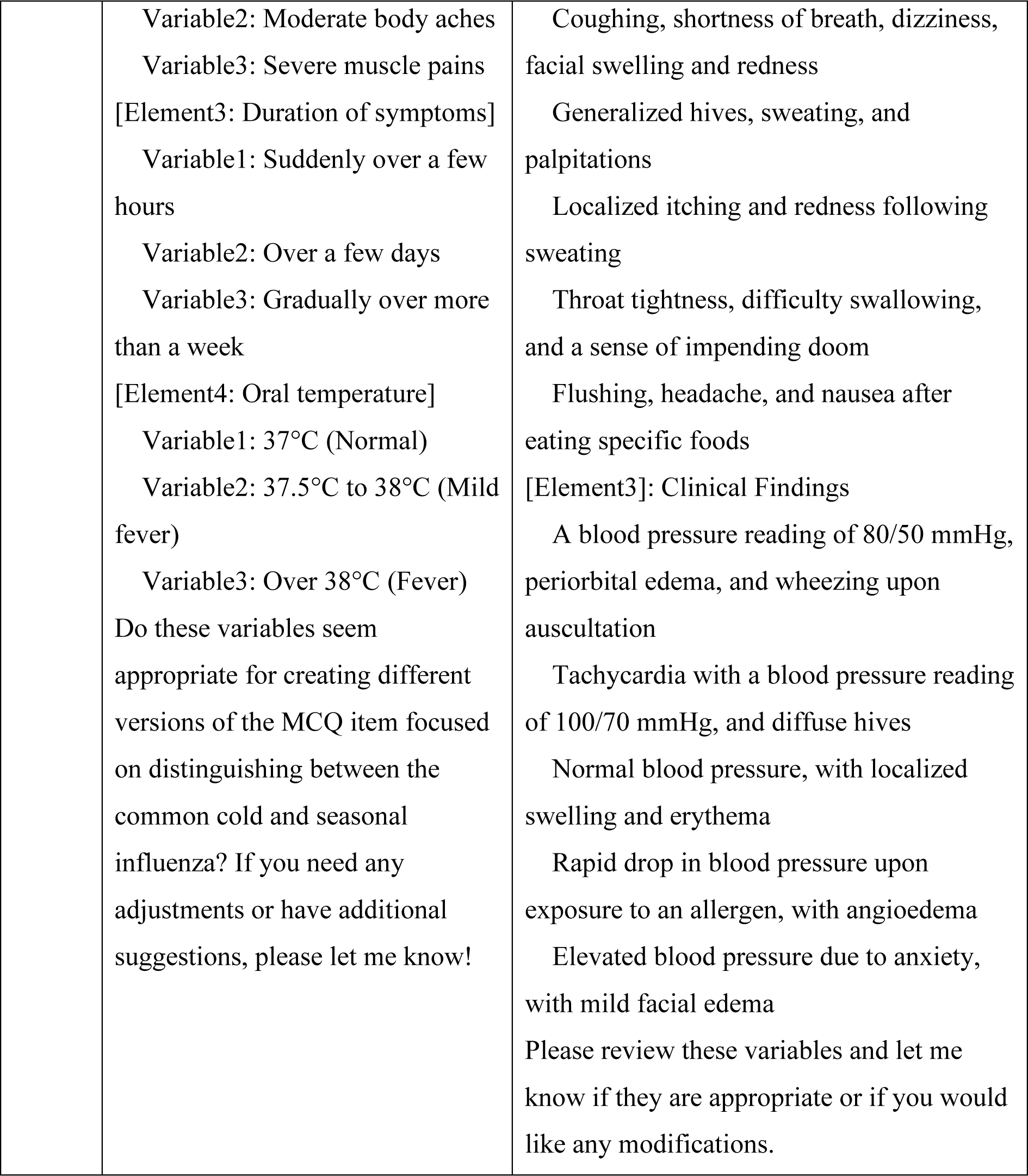
AI’s offer for values.

### Step 5: Generating the cognitive model

In this step, AI determines the constraints based on values (called “variables” by AI) approved by the expert, in order to create the cognitive model. Then, the expert reviews the output and prompts AI for necessary changes, if any. The presence of the expert is to validate the output, as it is in the previous steps.

Compared to the cognitive model in the traditional template-based method, it is more simple in our hybrid method. By providing constraints, it allows to determine what items will be generated and what the correct option is in each item. Table 4 and Table 5 present the cognitive models. Although the content needs small adjustments that can be easily carried out by the expert (e.g. the expert may ask ChatGPT to remove “37.5°C to 38°C” to make the correct answers more evident), we did not make any changes to present ChatGPT’s original output in order for demonstrating that it is able to provide a useful cognitive model to begin with for further revisions. It is also possible due to the nature of GPT models that the output might defer in formatting as visible in the tables.

**Table 4.** Cognitive model generated by AI for Gierl et al.’s item.

| **Element (Variable)** | **Constraints (Supported Options)** |
| --- | --- |
| **Cough** | Persistent dry cough (Seasonal Influenza) |
|  | Mild cough (Common Cold) |
|  | Severe hacking cough (Seasonal Influenza) |
| **Body aches** | Slight body aches (Common Cold) |
|  | Moderate body aches (Seasonal Influenza) |
|  | Severe muscle pains (Seasonal Influenza) |
| **Duration of symptoms** | Suddenly over a few hours (Seasonal Influenza) |
|  | Over a few days (Common Cold) |
|  | Gradually over more than a week (Common Cold) |
| **Oral temperature** | 37°C (Normal) (Common Cold) |
|  | 37.5°C to 38°C (Mild fever) (Seasonal Influenza) |
|  | Over 38°C (Fever) (Seasonal Influenza) |

**Table 5.**
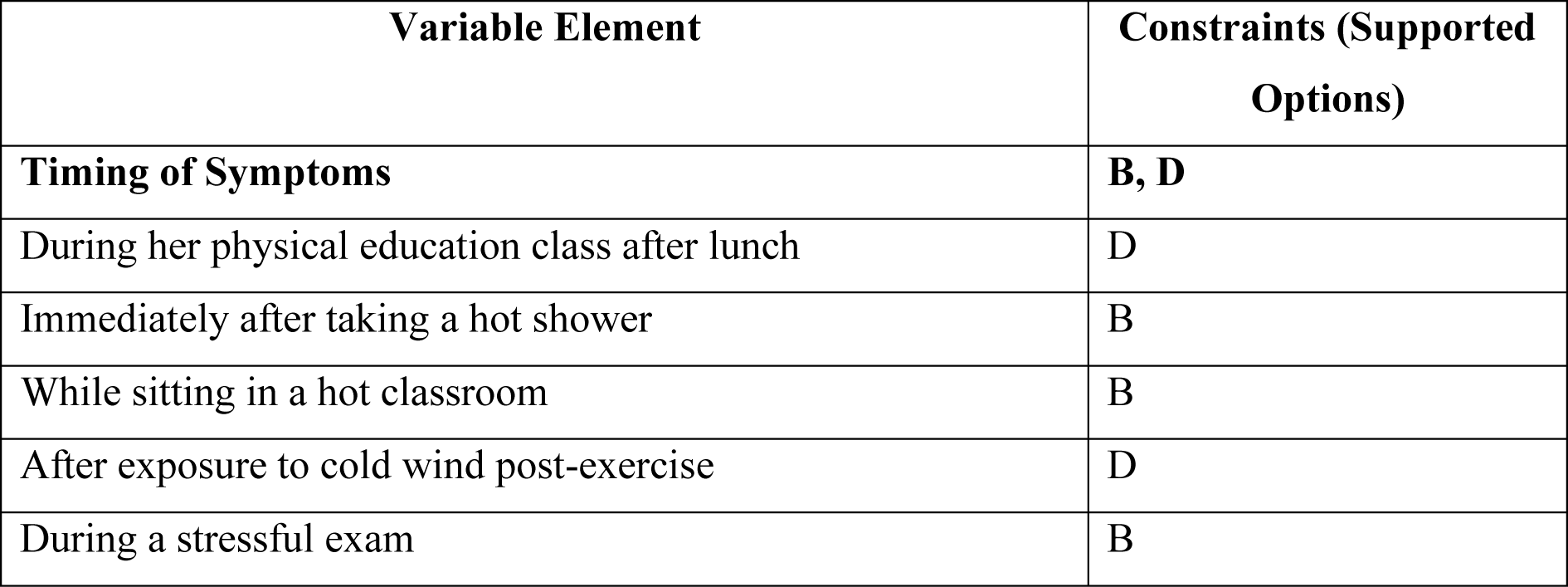
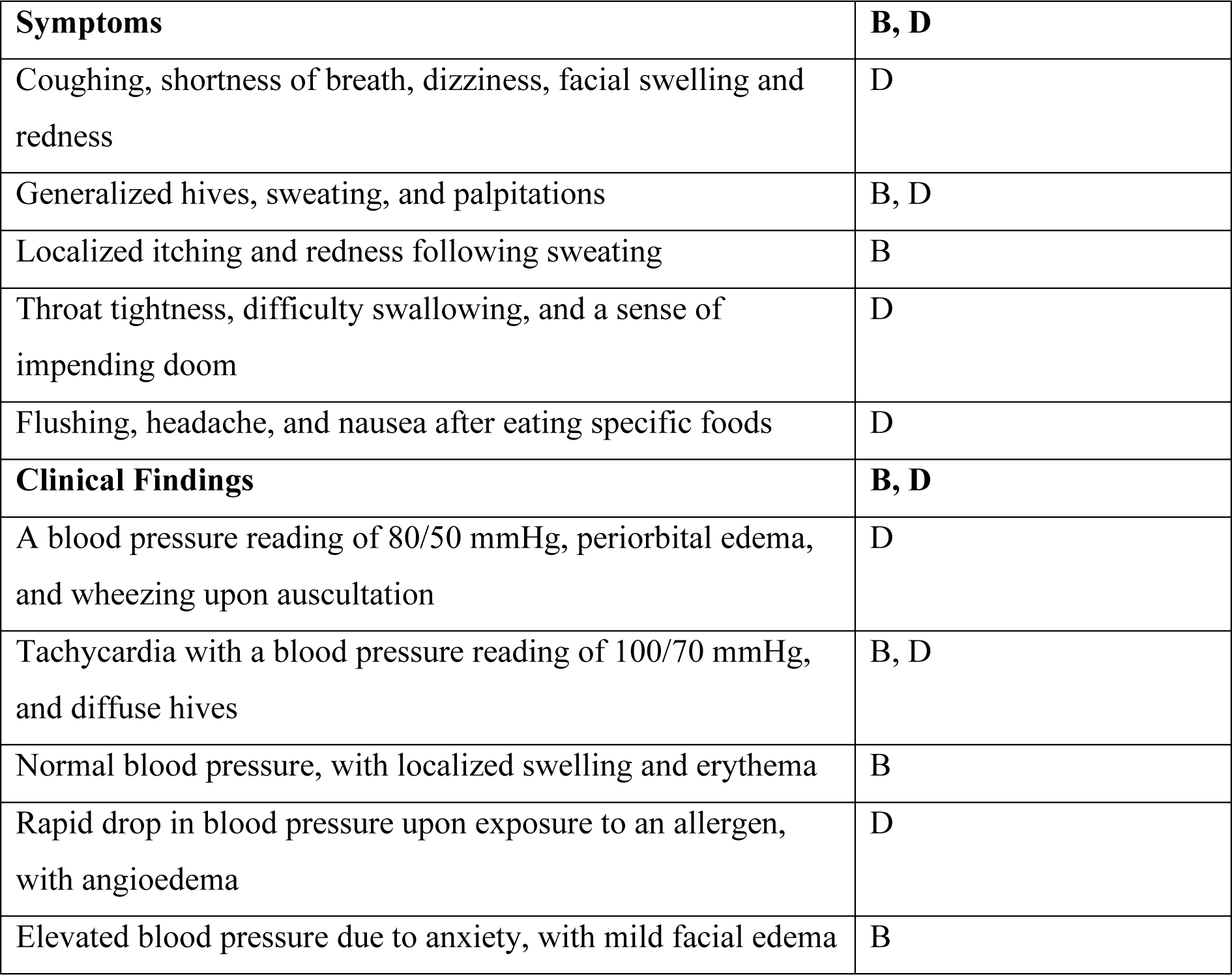
Cognitive model generated by AI for TUS item.

These five steps have been completed in less than 10 minutes for each model using a Custom GPT developed by us, titled *Item Model Maker for AIG*. The whole process within the AI environment can be displayed by accessing these public pages of the human-AI conversations: Gierl et al.’s item: https://chat.openai.com/share/1c975655-6bc4-4a16-a082-efad2030a5e2 TUS item: https://chat.openai.com/share/7e093fef-8d43-4b8f-bd15-a00bdf4c25c5

### Step 6: Final review by the expert(s)

In this step, the expert carries out a final review of the item model and cognitive model provided by AI, preferably with other experts.

In the previous steps, in order to keep the demonstration simple and due to the fact that there was not a significant inaccuracy in the AI-generated content, the expert did not demand any additional changes during the process in AI environment. But in the hybrid method, the five steps within the AI environment should be actively monitored by the expert, and if necessary, the expert should input prompts to make changes because AI is always prone to provide inaccurate content and deviate from providing a consistent template. Expert oversight, and involvement if necessary, is a strong necessity for creating high-quality item models and cognitive models.

Following the first five steps, which can be completed in less than 10 minutes, the expert should carry out one more round of review for the item model and the cognitive model generated through human-AI collaboration. It would be better if the expert conducts this review together with other experts to make sure that there is no inaccuracy, inconsistency, or inappropriate way of presentation. The expert should keep in mind that content generated by AI, in this case ChatGPT, is generated through a large language model, so it could “hallucinate”^25^ some inaccurate information and relationships that are harmful for the output quality. Apart from that, in this step, the expert may prefer to add more elements and variables, such as age and gender, in a way that does not change the correct answers, in order for increasing the number of the items.

### Step 7: Item generation using a non-AI software

In this step, the expert inputs the final version of the item template and the constraints to a traditional template-based AIG tool (software without AI involvement), and then the software algorithmically produces multiple-choice items based on the input provided by the expert. It is crucial to emphasize that the expert must input the content accurately because traditional software is not able to deal with inconsistent type of inputs, unlike the ability of AI in the previous steps. There is no difference between the traditional template-based method (stage 3)^6^ and our hybrid method (step 7) in this regard. As is in the template-based method,^6^ Hybrid AIG as well allows the software to generate hundreds of consistent items based on one item model.

## Discussion

In this study, we utilized AI to generate item models and cognitive models for generating multiple-choice items by using these models for template-based AIG. We demonstrated that AI is capable of providing AIG templates for this purpose under the guidance and oversight of human expert. Leveraging AI in template development has significantly reduced the time investment from five hours^6^ to less than 10 minutes, and provided a smoother experience for experts in this challenging^3(p30)^ task.

In our hybrid AIG method, cognitive work required to be carried out by experts in the past^6^ is now shared by AI. It switches the role of experts from “the creators of item-cognitive models from scratch” to “the reviewer of AI-generated content”, which brings an important efficiency to AIG without sacrificing consistency and accuracy. Our hybrid AIG method transcends the traditional template-based approach by marrying the “art” that comes from AI as a “black box”^21^ with the “science” of algorithmic generation^6^ under the oversight of expert as a “marriage registrar”. Practically, this balanced fusion under human guidance reduces the extraneous cognitive load^12^ on experts by allocating the burdensome tasks to AI in order for enhancing human efficiency and allowing them to concentrate on refining and validating the AI-generated content.

Similar to our approach, a recent study incorporated a large language model into the process of developing reading comprehension items.^26^ While addressing a critical issue in item development for a non-healthcare setting, its direct application to medical education is challenging due to the inherent complexities of health professions education. Furthermore, this approach integrates AI only into generating unique sentences based on rules imposed by experts, leaving the essential cognitive work dependent on expert input, which remains inefficient for medical education. In our hybrid model, we employ AI not only for generating unique sentences but also for development of item models and cognitive models as a whole, hence transforming the role of experts from the main “cognitive workers” to reviewers. This shift reduces cognitive effort for experts while maintaining their essential contribution for accurate and consistent items. Therefore, our findings diverge from an important conclusion drawn in the study on the successful generation of reading comprehension items.^26^ In that study, humans designed the cognitive model due to the necessity of testing with clear intentions to fulfill specific requirements and constraints. While we agree with the importance of clarity and constraints, we cannot fully subscribe to the interpretation that “it is neither possible nor desirable to create specifications and instructions using artificial intelligence”^26^. Our research demonstrated that even a minimal human oversight can be sufficient for utilizing AI in the creation of specifications and instructions, particularly in challenging domains such as medical education, which suggests even greater possibility for less complex tasks like reading comprehension. By dismissing the potential of AI in this regard by labeling it as “impossible”, humans might inadvertently limit AI’s capacity to enhance efficiency in cognitive work needed to be done. Thus, we propose leveraging AI more effectively rather than relegating it to a lesser role.

Our study has some limitations. Although the templates generated by AI showed promising results, replicability depends on the consistency of the AI model, which is GPT-4 in this case. While our study demonstrated that a hybrid AIG is possible, future research should explore this further by using different parent items across various settings to generate MCQs. As this is a proof-of-concept study, there is a lack of empirical evidence supporting the efficacy of the proposed Hybrid AIG method, no qualitative reviews to assess the generated items’ quality, and a lack of quantitative item analysis since the items were not tested on medical students.

However, it is still valuable because it has shown for the first time that generating plausible, and possibly useful, item templates using AI is possible in medical education. In the future studies, we are planning to generate items using these templates and investigate their effectiveness using qualitative and quantitative methods. Another limitation is that we generated simple templates. There are multi-layered templates for AIG,^3^ which require relatively complex structures, that might require from us to use different custom AIs for this purpose.

To conclude, the Hybrid AIG is a promising novel method that leverages AI in development of templates for template-based AIG that transforms the traditional role of experts from creators to reviewers. This shift can significantly reduce the cognitive burden on experts and streamline the item generation process while ensuring high-quality outcomes.

## Data Availability

Not applicable because no dataset was produced.

